# microRNA Biomarkers in Paediatric Infection Diagnostics. Bridging the Gap Between Evidence and Clinical Application: A Scoping Review

**DOI:** 10.1101/2025.04.11.25325664

**Authors:** Oenone Rodgers, Anna De Beer, Thomas Waterfield

## Abstract

**Objective:** This scoping review aims to assess the evidence regarding miRNA associations with paediatric bacterial and viral infections.

**Introduction:** Febrile children present a challenge in emergency care, often leading to unnecessary antibiotics due to difficulty distinguishing bacterial from viral infections. Current biomarkers lack specificity, contributing to diagnostic uncertainty and antimicrobial resistance. MicroRNAs (miRNAs), detectable in blood and responsive to disease, show promise as improved biomarkers, but their role in infection differentiation remains unclear. This scoping review aims to map known miRNA associations with paediatric infections and evaluates study methodologies to identify the best approaches for miRNA-based diagnostics.

**Inclusion criteria:** Studies reporting on children under 18 years of age with acute bacterial or viral infections will be included. Articles must focus on host miRNA biomarkers in biofluids. Exclusions include chronic infections, parasitic infections, fungal infections, sexually transmitted infections, animal models, in vitro, tissue samples, and in silico studies.

**Methods:** The databases to be searched will include MEDLINE and Web of Science with an additional for grey literature search via Google, Google Scholar, and open Theses restricted to the English language. Titles and abstracts will be screened, and eligible articles will undergo full-text review. The results of the search and study inclusion/exclusion process will be reported. Reasons for exclusion during the full text review are presented in the PRISMA-ScR flow diagram. Data will be extracted into a chart, analysed as percentages to assess consensus, and summarized in descriptive text with tables.

## Introduction

Febrile children pose a significant challenge to healthcare systems worldwide, with fever being the leading cause of Emergency Department (ED) visits(Hagedoorn et al., 2020). Antibiotic overprescription is a major concern in paediatrics, as many children receive precautionary treatment due to an increased risk of serious bacterial infection(Hagedoorn et al., 2020; van de Maat et al., 2019). The often-excessive use of antibiotics in children contributes to rising healthcare costs and escalating global antimicrobial resistance(Costelloe et al., 2010).

The primary challenge clinicians face is the differentiation of infection between bacterial and viral, and the related lack of certainty with regards to antibiotic prescriptions. Biomarkers used in current practice, including C-reactive protein (CRP) and Procalcitonin (PCT) (Stol et al., 2019), fail to sufficiently stratify febrile children at presentation to the ED(Van den Bruel & Thompson, 2014). This is due to the non-bacterial infection-specific nature of these biomarkers, meaning that their values can significantly increase with other conditions. The lack of a reliable biomarker leads to this uncertainty and the over prescribing of antibiotics ‘just in case’ of a serious bacterial infection being missed.

Researchers have attempted to identify novel single protein biomarkers, with limited success(Rodgers et al., 2024). The focus in paediatric biomarker discovery has now switched to biomarker panels that include proteins and other biomolecules such as RNA. In 2021, a systematic review of host-RNA signatures for paediatric infection diagnostics highlighted that RNA biomarkers may provide improved diagnostic value for paediatric infection(Buonsenso et al., 2021).

MicroRNAs (miRNAs) are non-invasively accessible via blood tests and may serve as alternatives to CRP and PCT. miRNA have also been shown to have rapid abundance changes in response to disease(Condrat et al., 2020; Metcalf, 2024) and exhibit excellent specificity(Saliminejad et al., 2019). These qualities make miRNA an attractive biomarker candidate to assess for paediatric infection diagnostics. The overall objective of this scoping review is to assess the extent of the literature relating to host miRNA associations with paediatric bacterial or viral infection. The specific aims include; identifying all miRNA with known associations to bacterial and viral infection in children, preferred study methods for miRNA identification and areas of consensus and best practice approaches for paediatric sample investigation.

### Review question

What are the associations of host-miRNA during bacterial and viral acute infection for children under 18 years of age in biofluid samples?

### Inclusion criteria

| Inclusion Criteria | Exclusion Criteria |
| --- | --- |
| **0-18 years** | Parasite, fungal or sexually transmitted infections |
| **Bacterial or Viral infection** | Cancer related miRNA |
| **Host miRNA** | Pathogen miRNA |
| **Biofluid samples** | Animal models, *in vitro*, tissue samples, *in silico* only studies |
| **Human** | Pregnancy associated miRNA/ foetus |
| **Acute infection** | Chronic infection |

### Participants

Participants under 18 years of age with suspected acute bacterial or viral infection.

### Concept

Articles need to focus on active acute infection of any severity, and the miRNA targets must originate from the host, not the pathogen. The focus of the study must be to measure within biofluids host miRNA responses to infection *in vivo*, and so articles only including animal models, *in vitro*, tissue samples or *in silico* will be excluded.

### Context

Any healthcare setting, primary or secondary care, globally where children are recruited with active acute infection.

### Types of sources

This scoping review will consider quantitative experimental study designs including randomised controlled trials, non-randomised controlled trials, before and after studies and interrupted time-series studies. In addition, systematic reviews that meet the inclusion criteria will also be considered, depending on the research question.

## Methods

The proposed scoping review will be conducted in accordance with the Joanna Briggs Institute methodology for scoping reviews(Peters et al., 2020). The Preferred Reporting Items for Systematic reviews and Meta-Analyses extension for Scoping Reviews (PRISMA-ScR) will be used.

### Search strategy

The search strategy will be adapted for each database and/or information source. The reference list of all included sources of evidence will be screened for additional studies. The databases to be searched include MEDLINE (PubMed) and Web of Science alongside searching for grey literature including Google, Google Scholar, and open Theses.

### Study/Source of evidence selection

Following the search, all identified citations will be collated and uploaded into Rayyan(Ouzzani et al., 2016) and duplicates removed. Titles and abstracts will then be screened by two independent reviewers for assessment against the inclusion criteria for the review. Reasons for exclusion of sources of evidence at full text that do not meet the inclusion criteria will be recorded and reported in the scoping review. Any disagreements that arise between the reviewers at each stage of the selection process will be resolved through discussion and a 3^rd^ reviewer a required. The results of the search and the study inclusion process will be reported in full in the final scoping review and presented in a PRISMA flow diagram(Tricco et al., 2018).

### Data extraction

The data extraction will be conducted by the lead author using a data extraction chart developed by the reviewers. The following items will be extracted: title, year of publication, authors, DOI, date of publication, country of study, population (e.g. children with fever), setting (e.g. emergency department, UK), clinical question (e.g. bacterial vs viral), main study methods, next-generation sequencing kit, RNA extraction kit, normalisation methods, ages of patients, % female, total sample size, sample size study focus, sample size comparator, bacterial numbers and infections, viral numbers and infections, inclusion of CRP/ PCT, reference standard, conditions of statistics, biomarkers, biomarker sensitivity %, biomarker specificity %, AUC, positive predictive value, negative predictive value, biomarker cut-off value, sample type, and sample volume.

A draft extraction form is provided (see Appendix 1). The draft data extraction chart will be modified and revised as necessary during the process of extracting data from each included evidence source. Modifications will be detailed in the scoping review. Any disagreements that arise between the reviewers will be resolved through discussion, or with an additional reviewer/s. If appropriate, authors of papers will be contacted to request missing or additional data, where required.

### Data analysis and presentation

The intention of the scoping review is to map and summarise available evidence, not to synthesise novel results. Therefore, data will be analysed as percentages of a whole to indicate the consensus from all the combined studies. The data gathered from the included studies will be discussed in a descriptive text and accompanied by tables.

## Data Availability

All data produced in the present work are contained in the manuscript.

## Author contributions

O.R conceptualised the research; O.R, A.D.B and T.W will act as reviewers; O.R writing—original draft preparation, O.R writing—review and editing; T.W. supervision; T.W funding acquisition. All authors will have read and agreed to the published version of the manuscript.

## Appendices

### Appendix I: Search strategy

#### MEDLINE all (Ovid)

miRNA.mp. MicroRNAs/ OR infection.mp. OR infections/ OR infectious disease.mp. OR bacterial infection.mp OR bacterial infections/ OR viral infection.mp OR virus diseases/ AND children.mp. OR child/ OR Infant/ or Adolescent/ or Pediatrics/ or paediatrics.mp. or Child, Preschool/ OR Neonatal Sepsis/ OR Intensive Care, Neonatal/ OR Intensive Care Units, Neonatal/ OR neonatal.mp.

#### Web of Science Core Collection

microrna* or miRNA* (All Fields) and infecti* or bacteri* or virus* or viral (All Fields) and child* or infant* or adolescent or p$ediatric* (All Fields) and Article (Document Types) and English (Languages)

### Appendix II: Data extraction instrument

|  | A | B | C | D |
| --- | --- | --- | --- | --- |
| 1 |  | Article 1 | Article 2 | Article 3 |
| 2 | title |  |  |  |
| 3 | year |  |  |  |
| 4 | authors |  |  |  |
| 5 | doi |  |  |  |
| 6 | date of publication |  |  |  |
| 7 | country of study |  |  |  |
| 8 | country |  |  |  |
| 9 | population (eg children with fever) |  |  |  |
| 10 | setting (eg ED, UK) |  |  |  |
| 11 | Clinical Question (eg bacterial vs viral) |  |  |  |
| 12 | study methods |  |  |  |
| 13 | Next-generation sequencing kit |  |  |  |
| 14 | RNA extraction kit |  |  |  |
| 15 | normalisation method |  |  |  |
| 16 | ages |  |  |  |
| 17 | % female |  |  |  |
| 18 | total sample size |  |  |  |
| 19 | Sample size Focus |  |  |  |
| 20 | Sample Sized comparator |  |  |  |
| 21 | Bacterial numbers and infections |  |  |  |
| 22 | Viral numbers and infections |  |  |  |
| 23 | CRP/ PCT |  |  |  |
| 24 | reference standard |  |  |  |
| 25 | Conditions of Sats |  |  |  |
| 26 | Biomarkers |  |  |  |
| 27 | sensitivity % |  |  |  |
| 28 | Specificity % |  |  |  |
| 29 | AUC |  |  |  |
| 30 | Positive predictive |  |  |  |
| 31 | negative predictive value |  |  |  |
| 32 | cut off value |  |  |  |
| 33 | sample type |  |  |  |
| 34 | sample volume |  |  |  |

